# Psychiatric disorders and self-harm across 26 adult cancers: cumulative burden, temporal variation, excess years of life lost and unnatural causes of deaths

**DOI:** 10.1101/2021.10.07.21264703

**Authors:** Wai Hoong Chang, Alvina G. Lai

**Affiliations:** Institute of Health Informatics, University College London, London, UK

**Author notes:** For correspondence: Alvina G. Lai.

**Keywords:** Cancer, psychiatric disorder, self-harm, suicide, cumulative burden, years of life lost

## Abstract

**Background:** Cancer is a life-altering event causing considerable psychological distress. However, population-representative variations in the total burden of psychiatric episodes across cancer types and treatment modalities have not been examined. We sought to estimate the risk of self-harm after incident psychiatric disorder diagnosis in patients with cancer, and the risk of unnatural deaths after self-harm.

**Design, Setting, Participants:** Population-based cohort study with multiphase study designs. Population-based linked patient records in England (1998-2020) from primary care practices, hospitals, cancer registry and death registry were employed. We identified 459,542 individuals age ≥ 18 years with an incident diagnosis of a site-specific cancer of interest.

**Main outcome measures:** Using outpatient and inpatient records, we identified patients with five psychiatric disorders of interest: depression, anxiety disorder, schizophrenia, bipolar disorder and personality disorder. Cumulative burden for all psychiatric events was estimated using the mean cumulative count method. We considered 10 cancer treatment regimens, 11 chemotherapy drug classes, deprivation status and 21 non-cancer comorbidities in stratified analyses. Propensity score matching was employed to identify controls who did not have any record of a psychiatric disorder of interest. For each psychiatric disorder category, we fitted a Cox regression model to estimate the risk of self-harm. We also estimated the risk of all-cause mortality and excess years of life lost comparing patients with and without psychiatric disorders. A separate matched cohort was generated to explore the risk of suicide and unnatural deaths following self-harm.

**Results:** Depression was the most common psychiatric disorder in patients with cancer, where some of the highest cumulative burdens were observed in patients with testicular cancer (98.05 per 100 individuals [CI: 83.08-127.25]), cervical cancer (78.74 [73.61-90.14]) and Hodgkin lymphoma (69.87 [61.05-69.48]) by age 60. Patients who received chemotherapy, radiotherapy and surgery had the highest cumulative burden of psychiatric disorders, while patients who received radiotherapy alone had the lowest burden. Patients treated with alkylating agent chemotherapeutics had the highest burden of psychiatric disorders while those treated with kinase inhibitors had the lowest burden. Among patients with cancer, 5,683 individuals were identified as having an incident self-harm episode. A previous diagnosis of psychiatric disorder before self-harm was at least twice as prevalent than a subsequent diagnosis of psychiatric disorder where the prevalence ratio was the highest in patients with brain tumours (5.36, CI: 4.57-6.14). Younger individuals were more likely to be diagnosed with mental illness before the first self-harm episode. However, individuals from more deprived regions (2.46, CI: 2.32-2.60) and individuals with ≥4 pre-existing comorbidities (2.19, CI: 1.92-2.46) were less likely to be diagnosed with mental illness before self-harm. Patients with mental illness had a higher cumulative burden of self-harm events compared with matched controls. All mental illnesses were associated with an increased risk of subsequent self-harm, where the highest risk was observed within 12 months of the mental illness diagnosis. Risks of self-harm during the first year in matched cohorts were as follow: depression (adjusted HR 44.1, CI: 34.0-57.1), anxiety disorder (HR 21.1, CI: 16.4-27.0) and schizophrenia (HR 7.5, CI: 5.0-11.2). Patients with cancer and psychiatric disorder experienced excess years of life lost. Patients who harmed themselves were 6.8 times more likely to die of unnatural causes of death compared with controls within 12 months of self-harm (HR 6.8, CI: 4.3-10.7). The risk of unnatural death after 12 months was markedly lower (HR 2.0, CI: 1.5-2.7).

**Conclusions:** This study quantifies the total burden of psychiatric events and self-harm in patients with cancer. The cumulative burden of psychiatric events varies across cancer type, treatment regimen and chemotherapy type. Incident psychiatric disorder diagnoses were significantly associated with increased risk of subsequent self-harm, where risks varied across psychiatric diagnostic categories and follow-up periods. Patients who harm themselves experienced the highest risk of dying from unnatural deaths within the first year of self-harm. We provide an extensive knowledge base to help inform collaborative cancer-psychiatric care initiatives by prioritising patients who are most at risk.

## Introduction

Mental illness is commonly found to be associated with an increased risk of mortality in patients with cancer^1–3^. Patients with cancer may experience significant psychological distress due to neuropsychiatric effects exerted by tumours, adverse reactions to physically demanding cancer treatment and substantial social and emotional impact (i.e., altered facial appearances) from cancer and its sequelae^4^. Cancer leaves permanent pathological alterations that imprint on people’s lives even when signs of active disease are no longer present. These effects, compounded by residual disability and periods of inability to work, could lead to social issues that serve to magnify psychological distress. Yet, cancer management often overshadows the recognition and treatment of psychiatric disorders. Patients with pre-existing mental health conditions may be prone to relapse during their cancer journey, while individuals without a history of mental health may face competing demands from their cancer that could distract physicians from recognising and diagnosing psychiatric disorders. Studies have also shown that individuals are often reluctant to seek professional mental health care^5^; of 24% of patients diagnosed with moderate or severe psychiatric disorder, only 8% had ever sought professional help^6^. Notable barriers to mental health help-seeking include a preference for self-reliance, low perceived need and negative experience or dissatisfaction with previous healthcare encounters^5^. Individuals may also be worried about being labelled as mentally ill and may experience self-blame, which further prompts catastrophic emotional and behavioural reactions against themselves (self-harm or suicide)^7,8^.

Systematic evidence of the total burden of psychiatric disorders, self-harm episodes and risk of suicide and unnatural deaths is essential to aid early identification and intervention of mental illness and suicidal thoughts. Detailed evidence is, however, lacking in this area. Real-world linked electronic health record data is uniquely well suited to address this question because: (1) it captures psychiatric and self-harm events in both community care and in-patient settings, (2) linkage to the cancer registry provide patient-level data on all cancer diagnoses and treatment regimens and (3) linkage to the death registry allows the complete ascertainment of cause of death. Our study seeks to: (1) estimate the variations in cumulative burden of five psychiatric disorders across 26 adult cancers stratified by treatment modalities and chemotherapy type, (2) estimate temporal variations in the first diagnosis of psychiatric disorder in relation to the time of the first self-harm event (prevalence ratios are calculated), (3) estimate the total burden of incident self-harm events (including recurrent events) after diagnosis of psychiatric disorder, (4) examine the subsequent risk of self-harm associated each psychiatric disorder at different timeframes, (5) examine mortality risk and excess years of life lost comparing patients with and without incident psychiatric disorder diagnosis and (6) investigate the risk of natural and unnatural deaths following self-harm. Our study identifies the timeframe and risk factors that can be used by physicians and family members to better monitor the mental health of patients with cancer. Psychiatric disorders are treatable modifiable risk factors, and efforts to recognise, diagnose and treat these conditions could positively affect the quality of life after cancer.

## Methods

### Linked electronic health record data sources

Two primary care linked electronic health record (EHR) databases, GOLD and Aurum, from the Clinical Practice Research Datalink (CPRD) were used. Information governance approval was obtained from the Medicines Healthcare Regulatory Authority Independent Scientific Advisory Committee (19_222 21_000593). The original dataset contains 5,343,578 individuals during the study period of 01-01-1998 to 31-10-2020. Primary care EHRs were linked to the secondary care Hospital Episode Statistics (HES), Office for National Statistics (ONS) death registry, patient-level Index of Multiple Deprivation (IMD; an area-based proxy for socioeconomic deprivation) and the National Cancer Registration and Analysis Service (NCRAS). Detailed cancer registration data (cancer site, behaviour, morphology and treatment information) were available in NCRAS. Chemotherapy drug details were obtained from the Systemic Anti-Cancer Treatment (SACT) dataset within NCRAS while radiotherapy details were obtained from the Radiotherapy (RTDS) dataset.

### Multiphase study designs

We identified incident primary site-specific cancer cases in individuals aged 18 years or older. Incident cancers were defined as the first diagnosis of cancer occurring at least 1 year after the start of the up to standard (UTS) date. The UTS date is a practice-based quality metric and an indication of research-quality patients and periods of quality data recording^9^. Patients had at least one year of UTS follow-up prior to the start of the study period (01-01-1998).

First, we analysed the cumulative burden of psychiatric disorders after cancer diagnosis. Since patients may experience recurrent psychiatric events, we employed the mean cumulative count method to not only capture the first occurrence of the event, but also subsequent occurrences. The cumulative burden method reflects a summarisation of all events that occur in a population by a given time^10^ (see cumulative burden section for details).

Second, we analysed temporal variations in the first psychiatric disorder diagnosis in relation to the first event of self-harm. The proportion of patients being diagnosed with a psychiatric disorder (for the first time) before and after the first self-harm event was calculated and plotted.

Third, we analysed the cumulative burden of recurrent self-harm events after the diagnosis of psychiatric disorders in patients with cancer.

Fourth, we estimated the risk of self-harm after diagnosis of psychiatric disorder in patients with cancer. We identified incident cases of psychiatric disorder (patients who had a history of psychiatric disorder before cancer diagnosis were excluded) (Figure S1). Patients with a history of self-harm before or at the time of the first diagnosis of psychiatric disorder were also excluded. For each psychiatric disorder, the first date of diagnosis (index date) was used as the date that follow-up started. Controls were identified by propensity score matching by age at cancer diagnosis, cancer type, sex, Index of Multiple Deprivation and primary care practice ID. Matching was performed using the optimal pair matching algorithm in the R matchit package based on the premise that the sum of the absolute pairwise distances in the matched sample was as small as possible. Unlike the nearest-neighbour matching, optimal matching ensures that within-pair distances remain small. For controls, the index date of its corresponding matched case was the date that follow-up started. Patients were followed up until self-harm, date of deregistration from the practice, death, or date of administrative censoring (31-10-2020), whichever occurred first (Figure S1).

Fifth, we estimated the risk of suicide and other causes of death following self-harm in patients with cancer. We identified incident self-harm episodes (patients who had a history of self-harm before cancer diagnosis were excluded). The first date of self-harm (index date) was used as the date that follow-up started. Controls were identified by propensity score matching by age at cancer diagnosis, cancer type, sex, Index of Multiple Deprivation and primary care practice ID. For controls, the index date of its corresponding matched case was the date that follow-up started. Patients were followed up until death, date of deregistration from the practice or date of administrative censoring (31-10-2020), whichever occurred first (Figure S2).

### Electronic health record phenotypes

All electronic health record (EHR) phenotypes were obtained from the open-access CALIBER phenotype library (https://portal.caliberresearch.org/) and have been previously validated^11–13^. Phenotypes for CPRD GOLD were generated using version 2 Read codes. Phenotypes for CPRD Aurum data were generated using a combination of SNOMED CT, Read version 2 and EMIS Web codes. Phenotypes for HES were generated in ICD-10. EHR phenotypes for self-harm were obtained from a previous study^14^. We employed primary care, secondary care and NCRAS records to identify patients aged ≥ 18 years with an incident primary site-specific cancer. We considered 26 cancer types: bladder, bone, brain, breast, cervix, colon and rectum, gallbladder and biliary tract, Hodgkin lymphoma, kidney and renal pelvis, leukaemia, liver and intrahepatic bile duct, lung and bronchus, melanoma, multiple myeloma, non-Hodgkin lymphoma, oesophagus, oropharynx, ovary, pancreas, prostate, small intestine, spinal cord and nervous system, stomach, testis, thyroid and uterus. We considered five psychiatric diagnostic categories: depression, anxiety disorders, schizophrenia, schizotypal and delusional disorders, bipolar affective disorder and mania and personality disorders.

Prevalent non-cancer physical comorbidities recorded before index date entry were identified from primary and secondary care records. We considered the following 21 comorbidities: heart failure, chronic obstructive pulmonary disease, HIV, hepatic disorders (i.e., alcoholic liver disease, non-alcoholic fatty liver disease, hepatic failure, liver fibrosis and cirrhosis and portal hypertension), stroke (i.e., ischaemic stroke, stroke not otherwise specified, transient ischaemic attack, intracerebral haemorrhage and subarachnoid haemorrhage), myocardial infarction, vascular disease (i.e., peripheral arterial disease, Raynaud’s syndrome, and venous thromboembolic disease) and abnormal glucose metabolism (i.e., type 1 and type 2 diabetes, diabetic neurological complications and diabetic ophthalmic complications). For stratified analyses, patients were divided into the following categories: 0 additional comorbidity (i.e., patients without diagnosis of any of the above non-cancer comorbidities), 1 additional comorbidity, 2 comorbidities, 3 comorbidities and 4+ comorbidities.

We analysed ten cancer treatment variables: (i) all chemotherapy (i.e., everyone who received chemotherapy), (ii) all radiotherapy, (iii) all surgery, (iv) chemotherapy only (i.e., individuals who received chemotherapy only and nothing else), (v) radiotherapy only, (vi) surgery only, (vii) chemotherapy and radiotherapy, (viii) chemotherapy and surgery, (ix) radiotherapy and surgery, (x) chemotherapy, radiotherapy and surgery. We considered 11 types of chemotherapy drug variables: (i) alkylating agents, (ii) anthracyclines, (iii) antimetabolites, (iv) biological response modifiers including monoclonal antibodies, (v) chemotherapy not otherwise specified, (vi) hormonal agents including corticosteroid hormones and sex hormones, (vii) kinase inhibitors, (viii) non-anthracycline antitumour antibiotics, (ix) plant alkaloids and natural products excluding vinca alkaloids, (x) platinum agents and (xi) vinca alkaloids. Cause-specific mortality was defined using the underlying cause of death code as recorded in the ONS death registry. Natural deaths consist of all ICD-10 codes, excluding V01-Y98. Unnatural deaths include V01-Y98 and suicides (including open verdicts X60-X84 and Y10-Y34)^14^.

### Statistical analyses

We estimated the cumulative burden of five psychiatric disorders and self-harm episodes using the previously described and validated mean cumulative count (MCC) method^10,15^. Given that psychiatric and self-harm episodes can recur, we employed the MCC method which considers recurrent events – cumulative incidence only considers the first occurrence of the event in each individual. We analysed the burden of recurrent events in the presence of competing risks. We considered death as a competing risk event as it precludes the occurrence of other events of interest. MCC can adopt any positive number, as it is an estimation of the mean count of events per individual within a given population (not the probability of developing the event). Cumulative burden of psychiatric disorders following cancer diagnosis was estimated as the average number of psychiatric events occurring at a given age. MCC of 0.5 at age 40 means that there is an average of 0.5 events occurring per individual at age 40 (i.e., 50 events per 100 individuals). Cumulative burden of self-harm following psychiatric disorder diagnosis was estimated as the average number of self-harm events occurring at a given time of follow-up (i.e., time from psychiatric disorder diagnosis to self-harm). MCC of 0.16 by 5 years of follow-up means that there is an average of 16 events per 100 individuals at 5 years. 95% confidence intervals (CIs) were generated using the bootstrap percentile method^10^.

We performed Cox proportional hazards regression to determine: 1) the risk of self-harm after psychiatric disorder diagnosis and 2) the risk of suicide and other causes of death following self-harm. Cox regression was performed on matched cohorts and further adjusted for non-cancer comorbidities, cancer treatment and the presence of additional psychiatric disorders. Proportional hazards assumption was evaluated using the Schoenfeld residuals. We also estimated the cumulative incidence for: 1) all-cause mortality after psychiatric disorder diagnosis and 2) all-cause and cause-specific mortality after self-harm. Analyses on specific causes of death were adjusted for the competing risk of dying from other causes. Years of life lost (YLL) refers to the number of years lost in patients with a given disease and was estimated using the R package lillies^16^, which was validated by other studies^17–19^. We calculated excess YLL as the average YLL (in years) that patients with psychiatric disorders experience from the time of diagnosis in excess to those experienced by the control population (i.e., individuals without psychiatric disorders) of the same age. Excess YLL was estimated based on the age of psychiatric disorder onset at ages 30, 40, 50, 60 and 70. Analysed were performed using R (3.6.3) with the following packages: tidyverse, tableone, lillies, reshape, splines, survival, survminer, etm, mstate, cmprsk, DataCombine, data.table, ggsci and pals.

### Patient and public involvement

No patients were involved in the implementation of the study as anonymised data was obtained from the Clinical Practice Research Datalink without direct contact with patients.

## Results

### Patients with testicular cancer had the highest burden of psychiatric disorders compared with other cancer types

Between 1998 and 2020, we identified 459,542 individuals age ≥ 18 years with an incident diagnosis of a site-specific cancer of interest. Patient characteristics were presented in Table S1. We analysed cumulative burden of five psychiatric disorders (depression, anxiety disorders, schizophrenia, schizotypal and delusional disorders, bipolar affective disorder and mania and personality disorders) following cancer diagnosis and compared results across 26 cancer diagnostic groups. Cumulative burden was the highest for depression, followed by anxiety disorders (Figure 1A, Table S2). At age 60, for example, cumulative burdens per 100 individuals for depression ranked from highest to lowest by cancer type were as follow: testis (98.05 [CI: 83.08-127.25]), cervix (78.74 [73.61-90.14]), Hodgkin lymphoma (69.87 [61.05-69.48]), spinal cord and nervous system (60.37 [33.8-63.46]), thyroid (52.69 [40.74-60.98]), bone (39.17 [38.06-51.09]), breast (34.35 [33.77-35.33]), melanoma (25.98 [25.02-27.23]), oropharynx (24 [21.4-27.61]), ovary (21.92 [19.96-23.22]), small intestine (18.08 [17.47-27.52]), kidney and renal pelvis (16.24 [13.51-18.27]), uterus (14.44 [13.36-16.66]), non-Hodgkin lymphoma (13.93 [12.64-14.28]), leukaemia (13.84 [12.03-14.74]), brain (11.93 [10.8-13.33]), multiple myeloma (8.5 [6.4-9.84]), colon and rectum (5.7 [5.35-6.09]), bladder (4.28 [4.19-5.32]), oesophagus (3.98 [3.66-3.98]), stomach (3.2 [2.69-3.38]), lung and bronchus (3.11 [2.58-3.42]), gallbladder and biliary tract (3.04 [2.47-3.68]), liver and intrahepatic bile duct (2.75 [2.47-3.65]) and pancreas (2.42 [1.94-3.42]), prostate (2.4 [2.15-2.67]) (Figure 1A, Table S2). Patients with testicular cancer had the highest burden across all psychiatric disorders -cumulative burdens at age 60 were as follow: depression (98.05 per 100 individuals [83.08-127.25]), anxiety (83.54 [78.03-90.59]), schizophrenia (8.24 [4.72-11.76]), personality disorders (5.42 [0.20-15.75]) and bipolar disorders (2.50 [1.59-3.13]).

**Figure 1.**
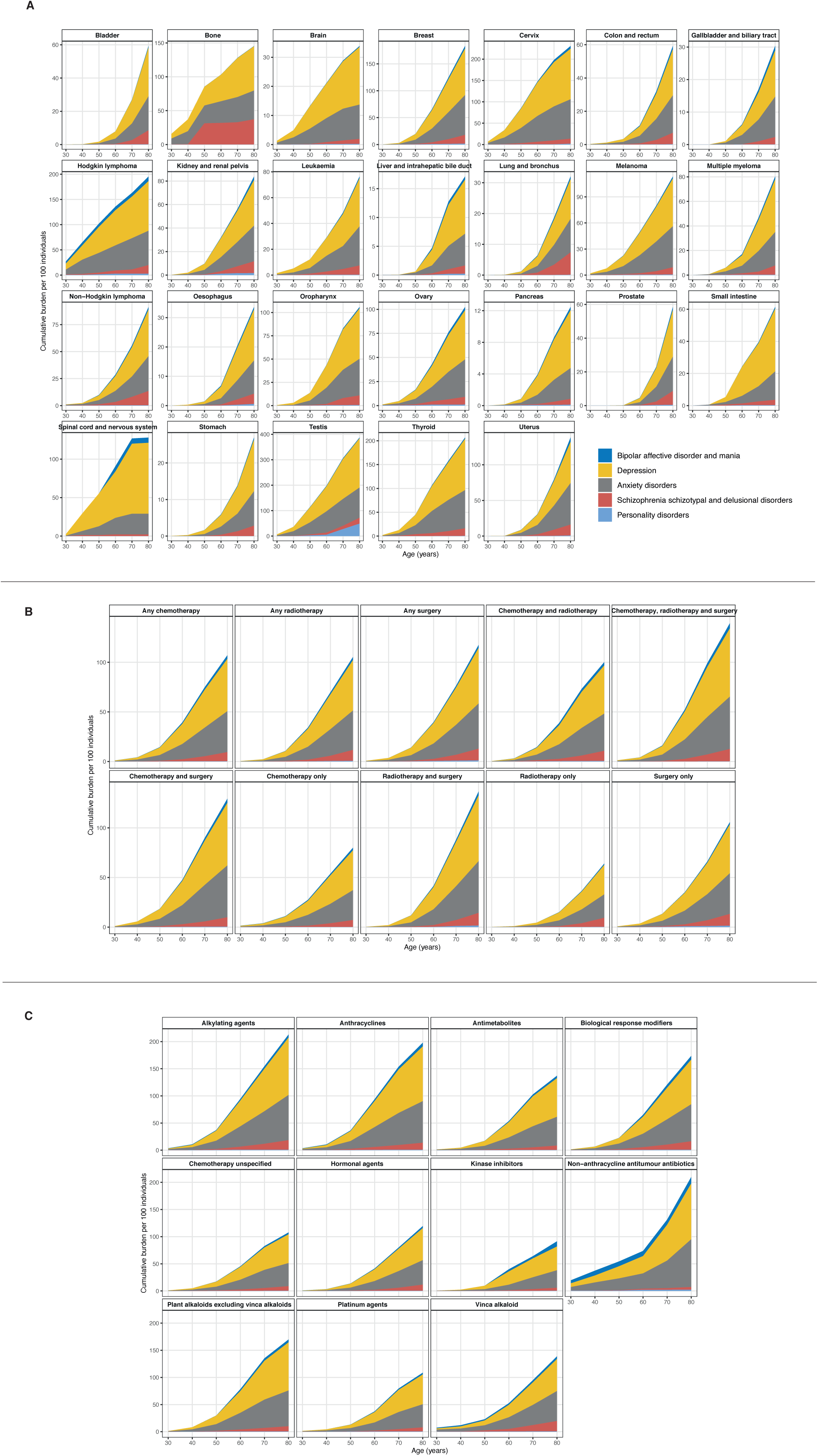
Cumulative burden of psychiatric disorders in patients with cancer. Distribution of cumulative burden by (A) 26 cancer diagnostic groups, (B) 10 cancer treatment modalities and (C) 11 chemotherapy drug class. All data and 95% confidence intervals are provided in the supplementary tables.

### Impact of cancer treatment on the burden of psychiatric disorders

When comparing across treatment modalities, patients who received all three chemotherapy, radiotherapy and surgery had the highest cumulative burden of psychiatric disorders (Figure 1B, Table S3). At age 60, cumulative burdens were: depression (28.23 per 100 individuals [27.07-29.94]), anxiety (19.66 [19.00-20.76]), schizophrenia (2.53 [2.10-2.96]), personality disorders (0.05 [0.02-0.07]) and bipolar disorders (1.94 [1.50-2.03]). At age 80, cumulative burdens were: depression (68.77 [67.17-71.07]), anxiety (52.70 [50.89-54.25]), schizophrenia (12.50 [11.83-13.75]), personality disorders (0.16 [0.13-0.20]) and bipolar disorders (5.18 [4.05-5.64]). By contrast, the lowest burden of psychiatric disorders was observed in patients who received radiotherapy alone. At age 60, cumulative burdens were: depression (8.09 [7.81-8.86]), anxiety (5.82 [5.55-6.60]), schizophrenia (0.80 [0.75-0.97]), personality disorders (0.09 [0.02-0.15]) and bipolar disorders (0.29 [0.20-0.46]).

### Patients who received alkylating agents for chemotherapy had the highest burden of psychiatric disorders

At age 60, cumulative burdens in patients who were treated with alkylating agents were: depression (47.55 per 100 individuals [45.31-51.21]), anxiety (37.47 [35.47-42.24]), schizophrenia (5.76 [4.84-6.18]), personality disorders (0.73 [0.60-0.97]) and bipolar disorders (3.14 [2.41-4.12]) (Figure 1C, Table S4). Patients who received kinase inhibitor treatment, by contrast, had the lowest burden of psychiatric disorders – cumulative burden at age 60 were: depression (24.46 [16.95-30.94]), anxiety (10.20 [6.85-11.93]), schizophrenia (1.14 [0.62-2.05]), personality disorders (0.14 [0.00-0.14]) and bipolar disorders (3.99 [0.05-4.03]).

### A diagnosis of previous psychiatric disorder was common in patients who self-harm

Among patients with cancer, 5,683 individuals were identified as having an incident self-harm episode (self-harm event after cancer diagnosis) (Figure S2). We estimated temporal variation in the first diagnosis of a psychiatric disorder in relation to the time of the first episode of self-harm. We observed that across cancer types, a previous diagnosis of psychiatric disorder prior to self-harm was at least twice as prevalent than a subsequent diagnosis of psychiatric disorder (Figure 2). Prevalence ratio was the highest in patients with brain tumours (5.36, CI: 4.57-6.14), followed by prostate cancer (4.30, CI: 4.08-4.52), Hodgkin lymphoma (4.17, CI: 2.98 -5.37), testicular cancer (3.96, CI: 2.87-5.04) and melanoma (3.84, CI: 3.44-4.24). By contrast, patients with lung cancer had the lowest prevalence ratio (2.05, CI: 1.90-2.20) (Figure 2A). Younger individuals were more likely to be diagnosed with mental illness before the first self-harm episode compared with older individuals. For example, individuals aged 18-34 were 4.3 times (CI: 3.69-5.06) more likely to be diagnosed with a psychiatric disorder prior to self-harm, compared with individuals aged 51-65 (2.66, CI: 2.54-2.77) (Figure 2B). Patients from the most deprived regions had a lower prevalence ratio (2.46, CI: 2.32-2.60) than those from least deprived regions (3.68, CI: 3.52-3.84), suggesting that socioeconomic deprivation had an impact on previous help-seeking behaviour before self-harm (Figure 2C). Similarly, patients with a high number of pre-existing conditions were less likely to be diagnosed with a psychiatric disorder before self-harm. Prevalence ratio for individuals with 0 non-cancer comorbidities was 3.24 (CI: 3.17-3.31) versus 2.19 (CI: 1.92-2.46) in individuals with 4+ comorbidities (Figure 2D).

**Figure 2.**
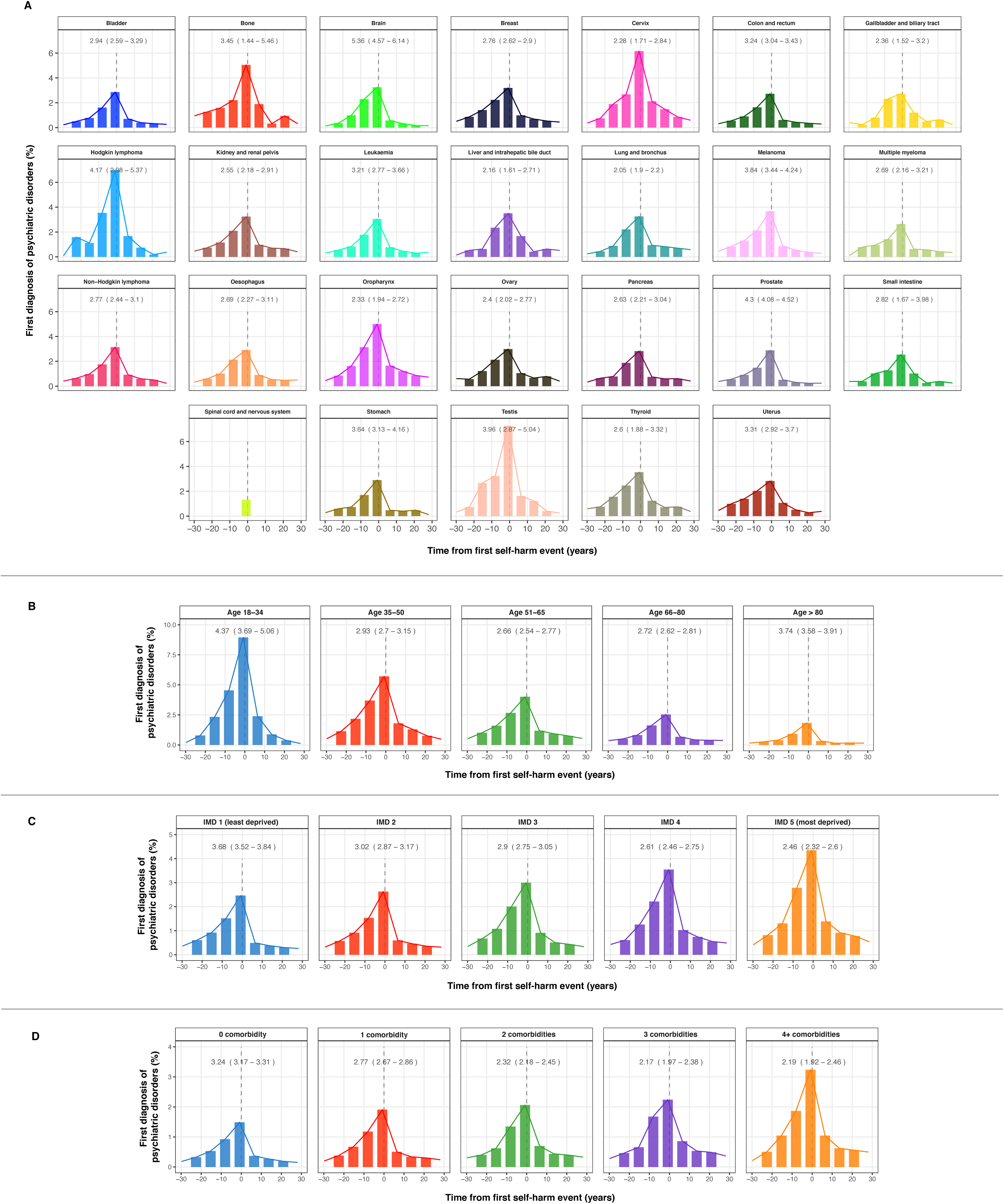
Temporal variation in the first diagnosis of psychiatric disorder in relation to the time of the first occurrence of self-harm. (A) By cancer type. (B) By age group. (C) By Index of Multiple Deprivation (IMD) status. (D) By the number of prevalent non-cancer comorbidities. Bars on the left side of the graphs indicate the proportion of individuals being diagnosed with a psychiatric disorder before the first self-harm event. Bars on the right side of the graphs indicate the proportion of individuals being diagnosed with a psychiatric disorder after the first self-harm event. Prevalence ratios (prevalence of psychiatric disorder diagnosis before self-harm divided by prevalence of psychiatric disorder after self-harm) were shown, alongside 95% confidence intervals.

### Total burden of non-fatal self-harm events and risk of self-harm following psychiatric disorder diagnosis

Propensity score matching was used to identify patients with and without a specific psychiatric disorder (Figure S1). The number of patients with incident psychiatric disorder were as follow: depression (21,609), anxiety disorder (20,070), schizophrenia (7,679), bipolar disorder (557) and personality disorder (194). The number of matched controls for each psychiatric disorder were as follow: depression (75,087), anxiety disorder (67,887), schizophrenia (23,130), bipolar disorder (1,636) and personality disorder (628) (Figure S1). Patient characteristics of the five diagnostic groups for cases and controls were summarised in Table S1. The cumulative burden of self-harm episodes was substantially higher in individuals with a prior diagnosis of psychiatric disorder compared. Across all control cohorts, the cumulative burdens of self-harm were less than 1 event per 100 controls at 10 years of follow-up, except for matched schizophrenia controls where the burden of self-harm was 1.36 events (CI: 1.16-1.52) (Figure 3A, Table S5). By contrast, cumulative burden of self-harm events per 100 individuals with mental illness at 1 year of follow-up were: depression (4.65, CI: 4.41-4.95), anxiety disorder (2.81, CI: 2.64-2.95), schizophrenia (1.16, CI: 0.95-1.44), personality disorder (7.75, CI: 4.65-9.29) and bipolar disorder (7.20, CI: 5.22-8.49). At 10-years of follow-up, the burden of self-harm were: depression (9.83 events per 100 individuals, CI: 9.73-10.45), anxiety disorder (7.93, CI: 7.81-8.35), schizophrenia (6.63, CI: 5.92-6.94), personality disorder (11.44, CI: 6.91-11.37) and bipolar disorder (43.88, CI: 40.72-49.56) (Figure 3A, Table S5).

**Figure 3.**
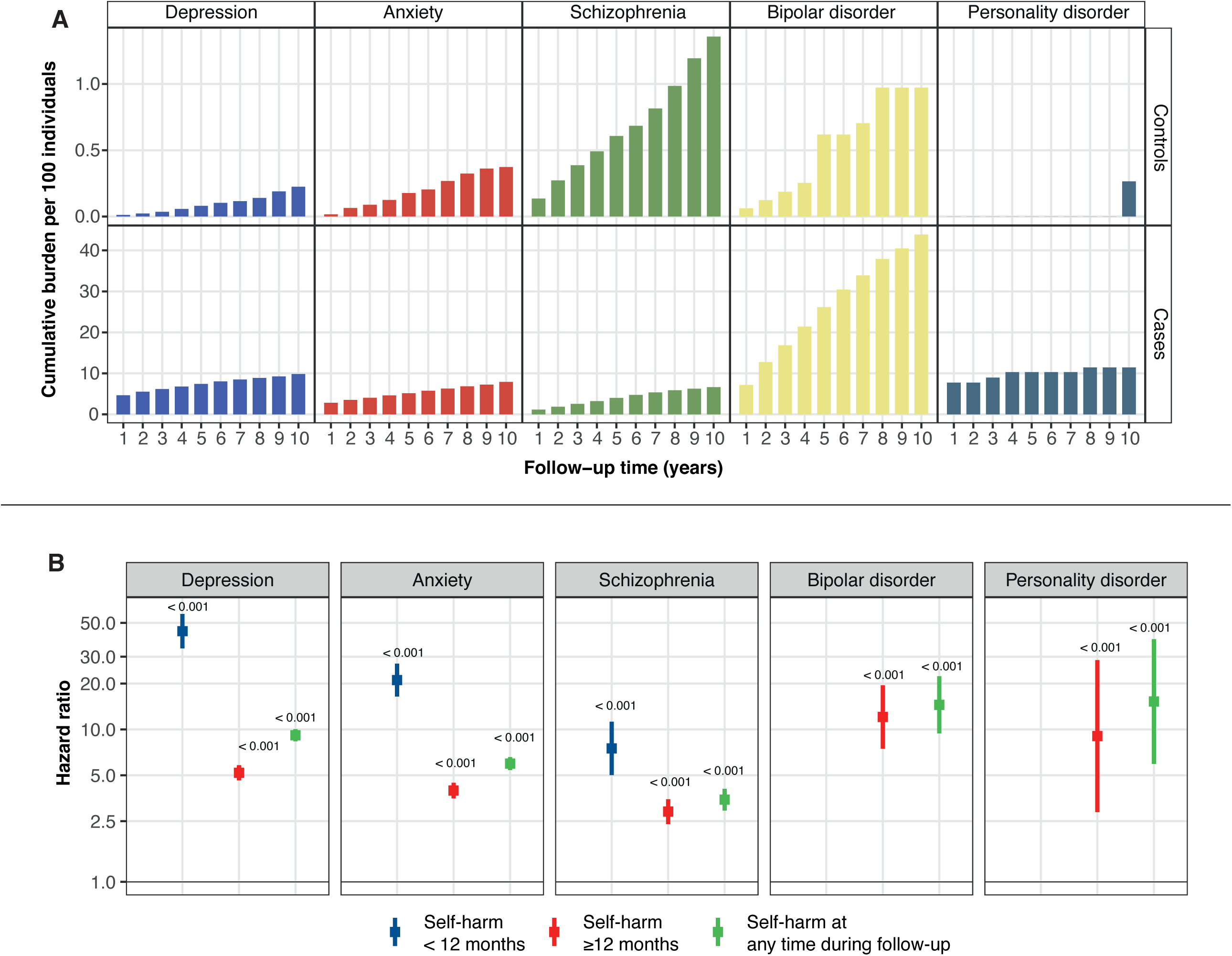
Total burden and risk of self-harm after diagnosis of psychiatric disorders in patients with cancer. Case (with psychiatric disorder) and control (no psychiatric disorder) groups were obtained via propensity score matching (see Figure S1). Controls were matched by age at cancer diagnosis, cancer type, sex, Index of Multiple Deprivation and primary care practice ID. (A) Cumulative burden of self-harm events in cases and controls. (B) Hazard ratios for risk of self-harm for each psychiatric disorder were further adjusted for non-cancer comorbidities, cancer treatment and presence of other psychiatric disorders. Self-harm risks during the first 12 months and subsequent years of follow-up were shown. Strata with low event numbers (n < 10) were not analysed. All data and 95% confidence intervals are provided in the supplementary tables.

All psychiatric disorders were significantly associated with an increased risk of subsequent self-harm (Figure 3B, Table S6). Self-harm risk was observed to change according to the length of follow up, with significantly higher hazard ratios (HRs) observed within 12 months of psychiatric disorder diagnosis. Individuals with depression were estimated to be 44 times more likely to self-harm during the first year (adjusted HR 44.1, CI: 34.0-57.1). Patients with anxiety disorder or schizophrenia were 21 times (HR 21.1, CI: 16.4-27.0) and 7 times (HR 7.5, CI: 5.0-11.2) more likely to self-harm, respectively, during the first year (Figure 3B, Table S6). The risk of self-harm markedly decreased over subsequent years of follow-up: depression (HR 5.2, CI: 4.6-5.8), anxiety disorder (HR 4.0, CI: 3.5-4.5), schizophrenia (HR 2.9, CI: 2.4-3.5), bipolar disorder (HR 12.1, CI: 7.5-19.5) and personality disorder (HR 9.0, CI: 2,9-28.5) (Figure 3B, Table S6).

### Patients with cancer and mental illness experienced excess years of life lost

Across all psychiatric disorders, the cumulative incidences for all-cause mortality were consistently higher in cases than controls (Figure 4A). We estimated excess years of life lost (YLL) which is the average number of years that patients with psychiatric disorder lose in excess of that found in patients without psychiatric disorder of the same age. Excess YLLs for each psychiatric disorder across different age of onset were displayed as radar plots (Figure 4B, Table S7). Younger age of psychiatric disorder onset was consistently associated with higher excess YLL. At age 30, patients with anxiety disorder lose 28.3 years in excess of their matched controls. By comparison, excess YLL in patients diagnosed with schizophrenia at age 30 was 32.2 years. At age 60, excess YLLs ranked from highest to lowest were as follow: personality disorder (16.0, CI: 13.8-18.9), schizophrenia (14.1, CI: 13.7-14.5), bipolar disorder (13.3, CI: 11.7-14.9), depression (12.9, CI: 12.6-13.1) and anxiety disorder (12.1, CI: 11.8-12.3) (Figure 4B, Table S7).

**Figure 4.**
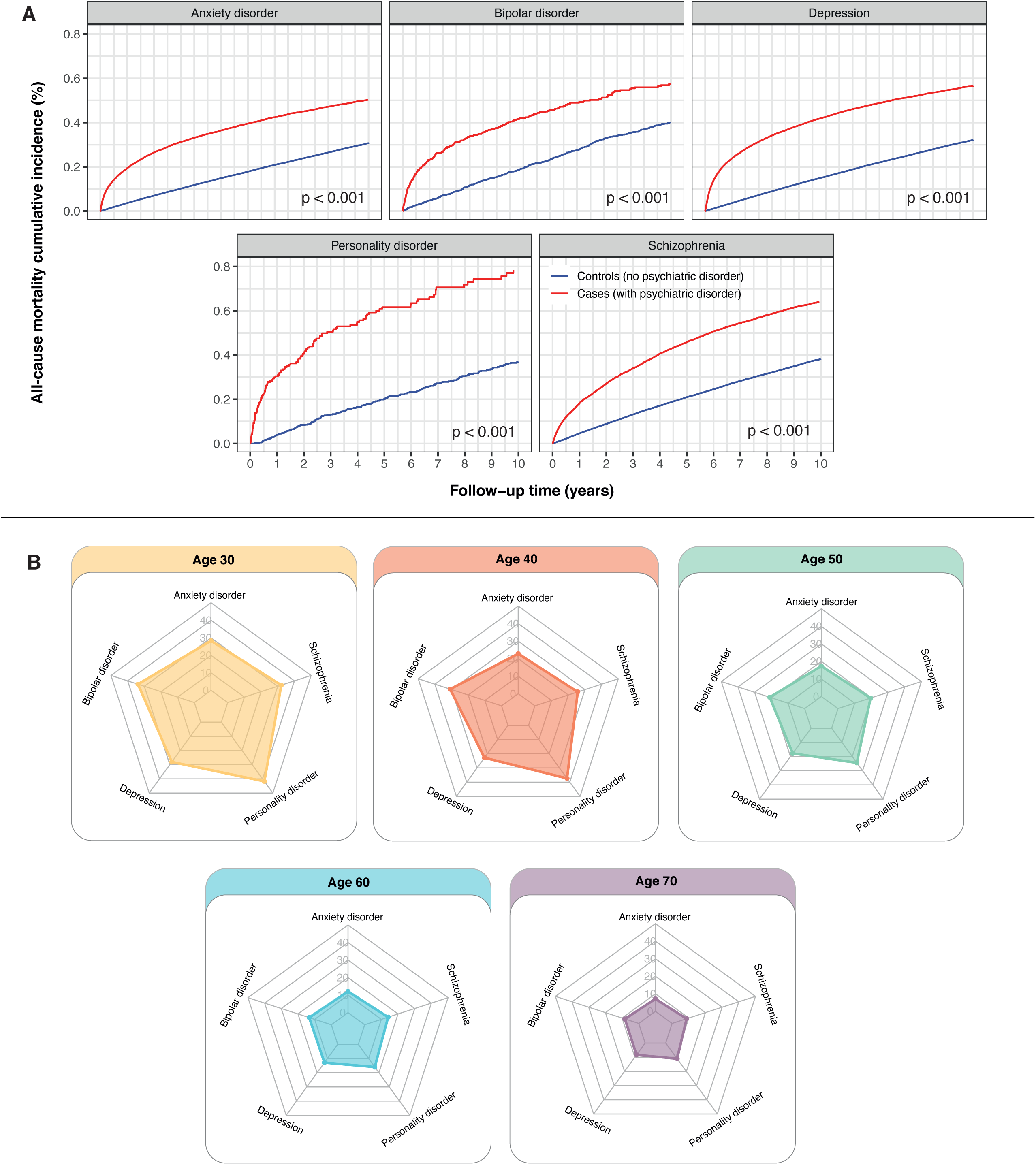
Cumulative incidence of all-cause mortality and years of life lost after diagnosis of psychiatric disorders in patients with cancer. Case (with psychiatric disorder) and control (no psychiatric disorder) groups were obtained via propensity score matching (see Figure S1). Controls were matched by age at cancer diagnosis, cancer type, sex, Index of Multiple Deprivation and primary care practice ID. (A) Cumulative incidence curves for all-cause mortality after psychiatric disorder diagnosis in matched case and control groups. (B) Excess years of life lost (YLL) attributable to psychiatric disorders in patients with cancer. Radar plots depict the difference in YLL between matched cases and controls. Excess YLL was estimated based on the age of onset of a psychiatric disorder. All data and 95% confidence intervals are provided in the supplementary tables.

### Patients who self-harm had the highest risk of unnatural deaths within 12 months of self-harm

Propensity score matching was used to identify cases (with self-harm) and controls (without self-harm) for analyses on cause-specific mortality risk following self-harm. We identified 5,683 individuals with incident self-harm episodes and 18,407 matched controls (Figure S2). Adults who harmed themselves were 6.8 times more likely to die of unnatural causes of death compared with controls within 12 months of self-harm (HR 6.8, CI: 4.3-10.7). The risk of unnatural death after 12 months was markedly lower (HR 2.0, CI: 1.5-2.7) (Figure 5A, Table S8). Cumulative incidence curves also demonstrated that the self-harm group had an increased risk of dying from unnatural deaths, after adjusting for competing risk of natural deaths (Figure 5B).

**Figure 5.**
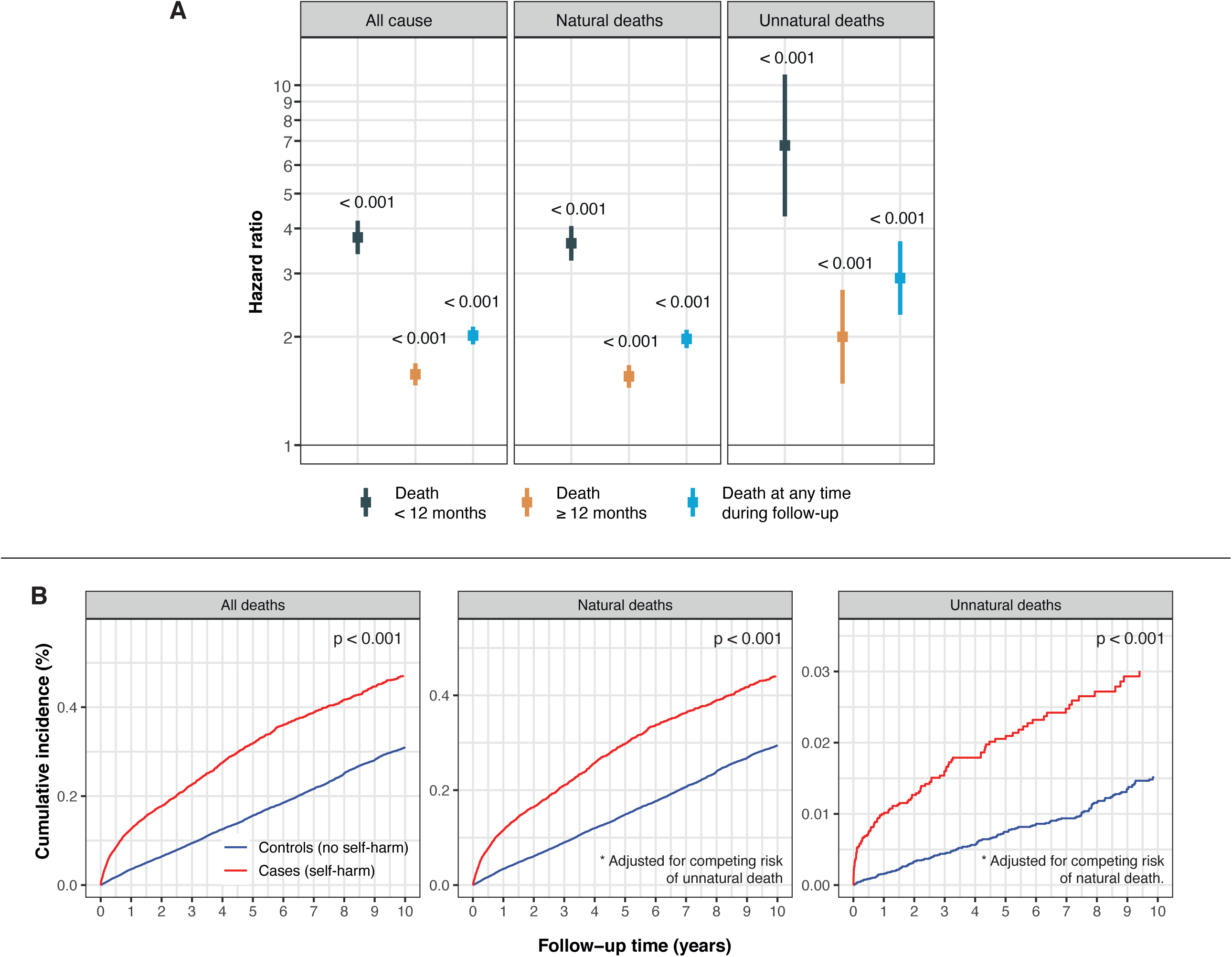
Risk of suicide and other causes of death following self-harm in patients with cancer. Case (self-harm) and control (no self-harm) groups were obtained via propensity score matching (see Figure S2). Controls were matched by age at cancer diagnosis, cancer type, sex, Index of Multiple Deprivation and primary care practice ID. (A) Hazard ratios for risk of suicide and other causes of death were further adjusted for non-cancer comorbidities, cancer treatment and presence of psychiatric disorders. Mortality risks during the first 12 months and subsequent years of follow-up were shown. (B) Cumulative incidence curves of death due to all causes, natural causes and unnatural causes after self-harm in matched case and control groups.

## Discussion

### Principal findings

We present the first study examining mental illness and self-harm events across 26 cancer types. Utilising data from primary care practices and hospitals, we quantified the total burden (not just the first event) of psychiatric disorder and self-harm. We also examined the prevalence of mental health diagnoses before and after self-harm and demonstrate that previous diagnoses of psychiatric disorders are important predictors of self-harm. We observed considerable differences in the risk of self-harm across psychiatric disorders. Patients with depression had the highest risk of self-harm, especially within 12 months of diagnosis, suggesting that patients require higher vigilance during this initial critical period. Interestingly, we found that schizophrenia was associated with a lower risk of self-harm – a finding that is corroborated by another study conducted in Hong Kong^20^. Patients with mental illness were significantly more likely to experience premature mortality. Furthermore, the risk of suicide and other causes of death were significantly higher in patients who harm themselves, particularly within 12 months of the first self-harm episode.

Two models have been proposed as contributing to the underlying cause of psychiatric disorders in patients with cancer: the biopsychosocial model and the neuropsychiatric effects of cancer and cancer treatment model^4^. The biopsychosocial model posits that biological, psychological and social factors contribute to chronic periods of psychological stress. Patients experiencing chemotherapy-induced alopecia are more likely to experience depression due to poorer body image and psychosocial well-being^21^. Patients may also experience health anxiety and fear of cancer recurrence, which can be triggered by internal (e.g., physical symptoms) and external cues (e.g., medical consultations, regrets about treatment decisions and media exposure)^22^. Coping with a life-threatening diagnosis of cancer may reinforce abnormal behaviours such as antisocial, narcissistic and obsessive-compulsive tendencies that define personality disorders^23^.

### Comparison with other studies

#### Neuropsychiatric effects of cancer

A meta-analysis found that the pooled mean prevalence of depression in patients with cancer ranged from 8% to 24%, which varies across cancer types and cancer treatments^24^. Similarly, another study demonstrated that 23% and 19% of patients with cancer experienced depression and anxiety, respectively where depression was more prevalent in patients in in-patient settings^25^. Patients with pancreatic cancer had higher levels of interleukin-6 cytokines, which is correlated with the severity of depressive symptoms^26^, but interestingly not with other measures of psychological distress. Patients with cancer may experience paraneoplastic neurologic syndromes, which are caused by immunological reactions to tumours. Paraneoplastic syndrome may induce psychiatric changes such as depression, personality disturbances, hallucinations and psychosis^27^. Patients experiencing paraneoplastic cerebellar degeneration exhibit psychiatric symptoms, for example, limbic encephalitis seen in patients with lung cancer presents with anxiety and depression^28^.

#### Neuropsychiatric effects of cancer treatment

The first presentation of bipolar disorder typically occurs at younger ages, however, recent studies found that 10% of elderly individuals develop the first onset of manic episodes later in life^29^. Patients with bipolar disorder experience recurring manic and depressive episodes, which can be exacerbated by cancer or its treatment. Although mania is less commonly seen in patients with cancer, it may be precipitated by steroids used as part of cancer treatment^30^ or mania secondary to brain cancer. Glucocorticoid steroid-induced manic and hypomanic symptoms are common, and symptoms are thought to be dose-dependent^31^. Another case report found that the chemotherapy 5-Fluorouracil induced manic episodes in a patient without a history of psychiatric illness^32^. Since 5-Fluorouracil could penetrate the blood-brain barrier, it is linked to neurotoxicity^33^ and mania may be caused by injury to neurotransmitter pathways. There have been limited studies on mania caused by extracerebral cancer and neuropsychiatric symptoms reported in these situations are either triggered by steroids or paraneoplastic syndromes due to antineuronal antibodies^34^. Two reports described first-onset mania in lung cancer and another demonstrates recurring mania in a patient with lung adenocarcinoma and a previous diagnosis of bipolar disorder^35,36^.

#### Impact of psychiatric disorders on cancer prognosis and subsequent self-harm behaviour

We have shown that psychiatric disorders have a major impact on life after cancer diagnosis where patients with psychiatric illnesses had a higher incidence of mortality and experienced excess years of life lost. Patients with schizophrenia are more likely to receive palliative care and experience premature mortality^37^, suggesting that disparities in health and cancer care exist and are influenced by the pervasive stigma of mental illness. High-intensity cancer care is associated with an increased risk of psychomotor agitation, paranoid delusions and recurrence of psychotic symptoms^38^. There has been very limited research exploring the risk of suicide and self-harm in patients with both cancer and schizophrenia. In general, patients with schizophrenia are 5 times more likely to commit suicide^39^. We also found that the risk of self-harm was the highest within 12 months of mental health diagnosis, suggesting that patients who are experiencing active psychological symptoms in earlier stages of the disease are more likely to harm themselves. In these high-risk patients, a lower threshold for psychiatric consultation and intervention may be beneficial to reduce self-harm risk. Patients with personality disorders are often rigid and inflexible and such behaviours may affect cancer progression either through the maintenance of an unhealthy lifestyle or the inability to cope with cancer, treatments and changes in life^23^. Patients with certain personality traits can feel alienated. Neuroticism is linked to poorer quality of life after cancer treatment in patients with breast cancer^40^. Neuroticism is also associated with long-term physical (e.g., peripheral neuropathy and tinnitus) and mental (poor self-esteem and unhealthy lifestyle) morbidities in patients with testicular cancer^41^. These patients may require additional support to help them better adapt to life with cancer.

### Strengths and limitations

First, this is the most comprehensive study examining psychiatric disorders across 26 adult cancers, excess mortality due to mental illness, risk of self-harm and risk of suicide and other causes of deaths using a single population-based cohort. The use of population-based data means that our findings are generalisable. Second, given that we have employed linked health records from primary care practices and hospitals, a particular strength is the ascertainment of all psychiatric and self-harm events in ambulatory and in-patient settings. Unlike in studies based on in-patient hospital records^20^, our results are less likely to be affected by biases due to under-hospitalisation or underdiagnosis of psychiatric disorders. We were able to capture mild cases managed in community primary care. Third, we relied on clinically recorded diagnoses of psychiatric disorders and self-harm episodes, which means that as opposed to self-reported events, our study is free of reporting biases. Fourth, we were able to examine the effects of cancer treatment type and chemotherapy type on subsequent psychiatric events as detailed information on cancer treatment is available from the cancer registry. Fifth, linkage to the national death registry enabled the analyses on cause-specific mortality with complete case ascertainment. Sixth, our study employs the mean cumulative count method to estimate the total burden of psychiatric and self-harm episodes over time. All other studies used the cumulative incidence method, which only considers the first event, thereby underestimating the total burden of recurring mental illness and self-harm.

We outline several limitations. While our use of population-based records provides robust and representative data, there remains a risk of underreporting of self-injurious behaviour due to stigma, particularly among more affluent communities. Our analyses were adjusted for socioeconomic deprivation to reduce the impact of these biases. Suicides may be underestimated as we have used ICD codes on death certificates as we do not have access to coroners’ reports. Coroners recording a suicide verdict must indicate suicidal intent beyond a reasonable doubt or else an open or accidental verdict is returned^42^. To address this problem, we have included open verdicts in our analyses as recommended by others^14,42^. We have not considered treatments for psychiatric disorders. The effects of psychiatric interventions on cancer survivorship can be explored in the future.

### Implications for policymakers, healthcare professionals, carers and patients

The variations in the burden of psychiatric disorders according to cancer diagnostic category, treatment type and chemotherapy type can help inform targeted prevention strategies aimed at high-risk groups. We outline three areas for consideration: 1) early recognition and treatment of psychiatric conditions and effective monitoring after self-harm episode, 2) collaborative psychiatric and cancer care and 3) managing cancer treatment-disruptive behaviours.

Psychiatric illness may present at any point in the cancer journey. A Danish study observed an increase in incidence rates of brain and lung cancers upon the first-time psychiatric in-patient or outpatient contact. As it is not likely that the psychiatric condition itself would cause an immediate and sudden increase in cancer risk, the authors concluded that psychiatric disorders may represent one of the earliest manifestations of cancer^43^. This suggests that screening for psychiatric symptoms in cancers with paraneoplastic potential may aid in early diagnosis and treatment of both cancer and psychiatric disorder. Slow-growing cancers such as meningiomas produce psychiatric symptoms before neurological symptoms become apparent^44^. An Evidence-Based Care guideline for managing depression in patients with cancer proposed eight specific recommendations^45^: (i) screening patients with cancer for depression, (ii) provide psychoeducation, destigmatise depression and investigate medical contributors to depression (i.e., vitamin B12, iron and folate levels and hypothyroidism), (iii) provide pharmacologic and psychological interventions, (iv) assess depression severity and follow stepped care approach, (v) consider collaborative care interventions involving oncologists, primary care practitioners and psychiatrists, (vi) referral to mental health specialists where there is a risk of self-harm, (vii) consider psychological therapies such as cognitive behavioural therapy and (viii) consider the use of antidepressant medication for severe depression.

Prescribing of antidepressants will need to take into account potential contraindications or drug interactions with cancer therapy. For example, the SSRI antidepressant Fluoxetine should be avoided in patients receiving tamoxifen treatment for breast cancer due to adverse drug interactions and increased risk of death^46^. A randomised trial (SMaRT Oncology-2) found that an integrated collaborative care model for depression in patients with cancer resulted in a better quality of life, health, reduced anxiety, pain and fatigue^47^. Patients subjected to the integrated cancer-depression care model received intensive therapy (antidepressant drugs and face-to-face psychological therapy), which highlights the importance of collaborative care approaches in achieving sustained treatment effects with a marginal increase in cost.

Disruptive behaviour in patients with psychiatric illness may interfere with cancer treatment and continuing care. Unlike mental health physicians, oncologists may not receive adequate training in dealing with behavioural problems and there has been limited guidance on managing clinical and legal risks associated with these clinically complex scenarios. Some institutes have developed policies to help physicians respond effectively to uncooperative and disruptive behaviour. For example, the main principles are to focus on problem behaviours in a non-punitive manner, introduce mental health consultation early in the cancer treatment pathway, design individualised responses to patient’s behaviour and set realistic expectations of behaviour^48^. Dealing with treatment-disruptive behaviour can be exhausting, hence, multidisciplinary support for the primary physician is crucial especially in the ambulatory oncology setting.

## Conclusions

Patients with both cancer and mental illness experience premature mortality and are at greater risk of self-harm. Gaining awareness of health disparities represents a significant step towards improving survival and well-being in the long term. Our work may inform new initiatives of integrated collaborative care to identify patients who are most at risk, to inform resource allocation, to identify patient and institutional barriers to implementation and to justify the delivery of a patient-centric model of care.

## Data Availability

All data produced in the present work are contained in the manuscript.

## Data sharing

The data used in this study are available on successful ethics application to the Clinical Practice Research Datalink (CPRD). All summarised data and results are made available as supplementary materials.

## Acknowledgements

This research uses data provided by patients and collected by the NHS as part of their routine care.

## Authors contribution

WHC and AGL conceived and designed the study. WHC undertook literature search. WHC and AGL analysed the data and interpreted the results. AGL supervised the research. WHC wrote the initial manuscript draft. AGL revised the manuscript draft. All authors read and approved the final manuscript.

